# National trends and operational drivers of vaccine wastage in Uganda, 2020–2025: a descriptive analysis of four tracer antigens

**DOI:** 10.64898/2026.06.17.26355922

**Authors:** Sharon Namasambi, Richard Migisha, Collins Ankunda, Nasif Matovu, Christine Lanyero, Paul Sagyiri, Emmanuel Okiror Okello, Deogratius Katongole, Patricia Eyu, Benon Kwesiga, Lilian Bulage, Yasiini Nuwamanya, Fred Nsubuga, Alex Riolexus Ario

## Abstract

**Background:** Vaccine wastage reduces immunisation efficiency, increases costs, and complicates supply forecasting. Uganda routinely monitors vaccine use, but national evidence comparing observed wastage with World Health Organization (WHO) and Uganda-specific planning thresholds has been limited. We described national and sub-national trends for four tracer antigens to inform supply-chain planning and forecasting.

**Methods:** We conducted a retrospective descriptive analysis of routinely reported immunisation data from Uganda’s District Health Information Software 2, 2020–2025. We analysed Bacille Calmette–Guérin (BCG), measles–rubella (MR), oral polio vaccine (OPV), and diphtheria–tetanus–pertussis-containing vaccine (DPT). Vaccine wastage was calculated as the proportion of issued doses not administered. Annual wastage rates were summarised using medians, and temporal trends were assessed using the Mann–Kendall test. Observed wastage was compared with WHO thresholds: BCG≤50%, MR≤25%, OPV≤10%, DPT≤15%, and Uganda’s planning thresholds: BCG≤70%, MR≤40%, OPV≤15%, DPT≤10%. Effective Vaccine Management reports were reviewed to summarise reported reasons for wastage.

**Results:** During 2020–2025, median national wastage was 40.6% for BCG, 25.9% for MR, 10.0% for OPV, and 9.2% for DPT. OPV wastage declined from 12.8% in 2020 to 8.0% in 2025, with a significant downward trend (τ_b_=−1.00; p=0.008). OPV and DPT wastage remained largely within their respective Uganda in-country thresholds (≤15% and ≤10%) for most of the study period, while BCG generally remained below the WHO threshold (≤50%) and MR frequently exceeded the WHO threshold (≤25%) but remained within Uganda’s planning threshold (≤40%) in most years. The proportion of districts exceeding both WHO and Uganda thresholds declined for OPV from 36.3% to 5.5% (p=0.024) and for DPT from 22.6% to 1.4% (p=0.013). Wastage was consistently higher in lower-level (Health Centre II and III) facilities, compared to hospitals. Among 50 service delivery points, reported reasons included low session attendance (66%), multi-dose vial policy non-compliance (28%), and vaccine expiry (12%).

**Conclusion:** Uganda achieved reductions in OPV wastage and district-level improvements in DPT wastage, while BCG and MR remained more variable and frequently had higher wastage. Strengthening adherence to the multi-dose vial policy and improving session planning at lower-level facilities could strengthen vaccine utilisation and forecasting.

**What is already known on this topic:**

- Vaccine wastage is a key indicator of immunisation programme efficiency and supply-chain performance.
- Lyophilised multi-dose vaccines usually have higher wastage than liquid vaccines because opened vials must be discarded soon after reconstitution.
- National evidence on vaccine wastage trends in Uganda has been limited.

**What this study adds:**

- Using national data from 2020 to 2025, this study describes vaccine wastage trends for four tracer antigens across all districts in Uganda.
- OPV wastage improved significantly, and district-level threshold exceedance declined for OPV and DPT.
- BCG and measles-rubella wastage remained more variable and higher in lower-level facilities.

**How this study might affect research, practice or policy:**

- The findings support stronger multi-dose vial policy implementation, better session planning, and improved routine wastage monitoring.
- They also suggest a need to review service-delivery approaches for lyophilised vaccines in low-volume settings.
- This evidence can help inform vaccine forecasting and supply-chain planning in Uganda and similar settings.

## Introduction

Vaccine wastage is a major concern for immunisation programmes because it means that some vaccine doses are issued but not used. This can increase programme costs, reduce supply efficiency, and make vaccine forecasting less accurate (1–6). Some wastage is expected, especially when vaccines are supplied in multi-dose vials and only a few children attend an immunisation session (3,5,7). However, high wastage may point to problems with session planning, stock management, cold-chain handling, documentation, or use of the multi-dose vial policy (3,7–9). The problem is greater for vaccines such as Bacille Calmette–Guérin (BCG) and measles–rubella (MR), which must be discarded soon after reconstitution (10). In contrast, liquid vaccines such as oral polio vaccine (OPV) and diphtheria–tetanus–pertussis-containing vaccine (DPT) can sometimes be kept for later use if multi-dose vial policy requirements are met (7,10,11). For Uganda’s immunisation programme, reducing avoidable wastage is important for improving vaccine availability, reducing unnecessary costs, and strengthening routine immunisation services (12,13).

To improve vaccine use, programme managers need reliable data showing where wastage is high, which vaccines are most affected, and whether wastage is within expected limits. WHO provides vaccine-specific wastage thresholds, and Uganda also uses its own planning thresholds for vaccine forecasting and procurement (3,4,12,13). However, Uganda has limited national evidence showing how vaccine wastage has changed over time, how it varies across districts and facility levels, and how observed wastage compares with both WHO and Uganda-specific thresholds. Most previous studies in Uganda have focused on selected districts or short periods, making it difficult to guide national planning (14,15). In addition, routine wastage figures alone do not explain why wastage occurs. Programme data need to be combined with operational reports to understand whether wastage is linked to low client turnout, poor vial labelling, expiry, cold-chain problems, or stock documentation gaps (9,14,15).

Given the need to improve vaccine utilisation, forecasting, and supply-chain efficiency in Uganda, national evidence is needed to show which vaccines, districts, and facility levels experience the greatest wastage and whether observed wastage exceeds expected thresholds. To address this gap, we analysed routine immunisation data from Uganda’s District Health Information Software 2 for all districts from 2020 to 2025, focusing on four tracer vaccines: BCG, MR, OPV, and DPT. We also reviewed Effective Vaccine Management reports to summarise reported operational reasons for wastage. Specifically, this study aimed to describe national, district-level, and facility-level patterns of vaccine wastage for BCG, MR, OPV, and DPT in Uganda during 2020–2025; compare observed wastage with WHO and Uganda-specific thresholds; and describe reported operational reasons for wastage.

## Methods and Materials

### Ethics statement

Approval to access and use DHIS2 and UNEPI supplementary data was obtained from the Ministry of Health (MOH). In addition, a non-research determination clearance was approved by the U.S. Centers for Disease Control and Prevention (CDC). The investigation was conducted in accordance with applicable U.S. federal regulations governing public health surveillance and data protection (see, for example, 45 C.F.R. part 46; 21 C.F.R. part 56; 42 U.S.C. §241(d); 5U.S.C. §552a; 44 U.S.C. §3501 et seq.). As this analysis involved routine program and surveillance data and posed no more than minimal risk to participants, written informed consent was not required.

### Study design

We conducted a retrospective descriptive analysis of routinely reported immunisation data extracted from the District Health Information Software 2 (DHIS2) platform for Uganda, covering the period January 2020 to December 2025. The analysis included data from all public and private-not-for-profit health facilities reporting immunisation services nationwide. DHIS2 serves as Uganda’s national Health Management Information System (HMIS) and is the primary platform for monitoring immunisation performance, vaccine utilization, and wastage indicators at all levels of the health system.

### Study setting

Uganda’s immunisation programme is coordinated by the Uganda National Expanded Programme on Immunisation (UNEPI) within the Ministry of Health. Immunisation services are delivered through fixed facility-based sessions and outreach services across a tiered health system. For this analysis, lower-level facilities were defined as Health Centre II and Health Centre III facilities, which provide primary care and usually serve smaller catchment populations. Higher-level facilities were defined as Health Centre IVs, general hospitals, regional referral hospitals, and national referral hospitals, which generally serve larger catchment populations and provide broader clinical services. Vaccines are procured centrally and distributed through the National Medical Stores to district vaccine stores, from which they are supplied to health facilities. District health teams oversee vaccine distribution, cold-chain management, supportive supervision, and routine reporting.

### Tracer antigens

Tracer antigens were defined as core vaccines routinely monitored to assess immunisation programme performance, supply chain efficiency, and system continuity (16). Four antigens were selected: BCG, MR, OPV, and DPT, representing both lyophilised and liquid formulations with different Multi-Dose Vial Policy (MDVP) eligibility (2,17). BCG and MR are lyophilised (freeze-dried) vaccines without preservatives in 20 and 10 dose vials respectively, while OPV and DPT are preservative containing liquid vaccines in 20 and 10 dose vials, administered orally or by injection according to schedule.

### Data sources and collection

For each tracer antigen, annual facility- and district-level data were extracted from District Health Information Software 2 (DHIS2), including total vaccine receipts, opening and closing stock balances, and total doses administered to eligible individuals for each reporting period.

### Definition and calculation of vaccine wastage

We defined vaccine wastage as the proportion of vaccine doses that we discarded, damaged or destroyed, expressed as a percentage, in accordance with guidance from WHO (3,4).

We calculated wastage rates using the standard formula:

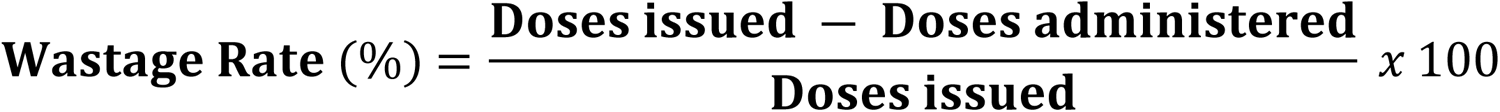

Where Doses issued = (doses received from higher-level stores + opening balance) – (closing balance), and

Doses administered = doses given to eligible individuals as recorded in DHIS2.

### Thresholds comparison

Observed vaccine wastage was assessed against two reference frameworks: WHO normative thresholds, representing global performance benchmarks, and Uganda’s in-country anticipated thresholds, used for vaccine quantification and procurement. WHO thresholds were BCG ≤50%, MR ≤25%, OPV ≤10%, and DPT ≤15%, while Uganda’s anticipated thresholds were BCG ≤70%, MR ≤40%, OPV ≤15%, and DPT ≤10%. For BCG, MR, and OPV, district-year observations were classified into three categories: within the WHO threshold, above the WHO threshold but within Uganda’s planning threshold, and above both WHO and Uganda thresholds. For DPT, where Uganda’s planning threshold (≤10%) is more stringent than the WHO threshold (≤15%), district-year observations were interpreted separately as within both thresholds, above Uganda’s threshold but within the WHO threshold, or above both thresholds.

### Data quality assessment

We assessed data quality before analysis. Reporting completeness was reviewed by district, facility level, antigen, and year. Facility-years with reporting completeness below 80% were flagged and described but retained in the main descriptive analysis to preserve national programme coverage. We also conducted range and internal consistency checks. Wastage values below 0% or above 100% were flagged and cross-checked against opening balance, doses received, closing balance, and doses administered. Stock-balance consistency was assessed by comparing reported closing balance with the expected closing balance calculated from opening balance, doses received, and doses administered. Implausible or inconsistent records like those with zero or negative issued doses were excluded from wastage-rate.

### Data management and analysis

Data were cleaned and analysed using Stata version 17 (StataCorp, College Station, TX, USA). Annual vaccine wastage rates were summarised at national, district, and facility category levels (lower levels vs. higher levels) for each antigen. For each antigen, annual national wastage rates were first calculated using aggregated national stock and administered-dose data for each year; the reported median national wastage represents the median of these six annual national estimates. We assessed temporal trends in national median wastage rates using the Mann–Kendall test, a non-parametric method appropriate for monotonic trend detection in short, non-normally distributed time series. The test yields Kendall’s tau-b (τ_b_), which ranges from −1 to +1, where negative values indicate a declining trend and positive values an increasing trend. An associated p-value indicating whether the observed trend is statistically significant was set at p ≤ 0.05; however, given the short six-year time series and the operational nature of the data, greater emphasis was placed on trend direction and magnitude rather than statistical significance alone. Annual percentage change was calculated descriptively to characterise year-to-year variation.

Spatial patterns were examined using district level choropleth maps generated in QGIS version 3.42.1. Districts were classified by threshold performance to identify geographic variation. Reported reasons for vaccine wastage from Effective Vaccine Management reports were grouped into operational factors, supply-chain factors, and documentation-related factors. These findings were summarised using frequencies and percentages.

Because EVM assessments were not designed as a representative causal study of wastage determinants, they were used to contextualise DHIS2 findings rather than to estimate population-level causes of wastage.

## Results

### Annual median vaccine wastage rates, 2020-2025

Between 2020 and 2025, national BCG wastage had a median of 40.6% (IQR 39.9–44.6), generally below the WHO threshold ≤50% but with variability from 37.6% to 72.5%.

MR wastage had a median of 25.9% (IQR 17.3–30.3), frequently exceeding the WHO threshold ≤25% but remaining within the national threshold ≤40% in most years. DPT median wastage was 9.2% (IQR 7.7–11.5) with moderate fluctuation. OPV wastage declined from 12.8% in 2020 to 8.0% in 2025, with low variability (median 10.0%; IQR 9.4–10.7) (Figure 1).

**Figure 1:**
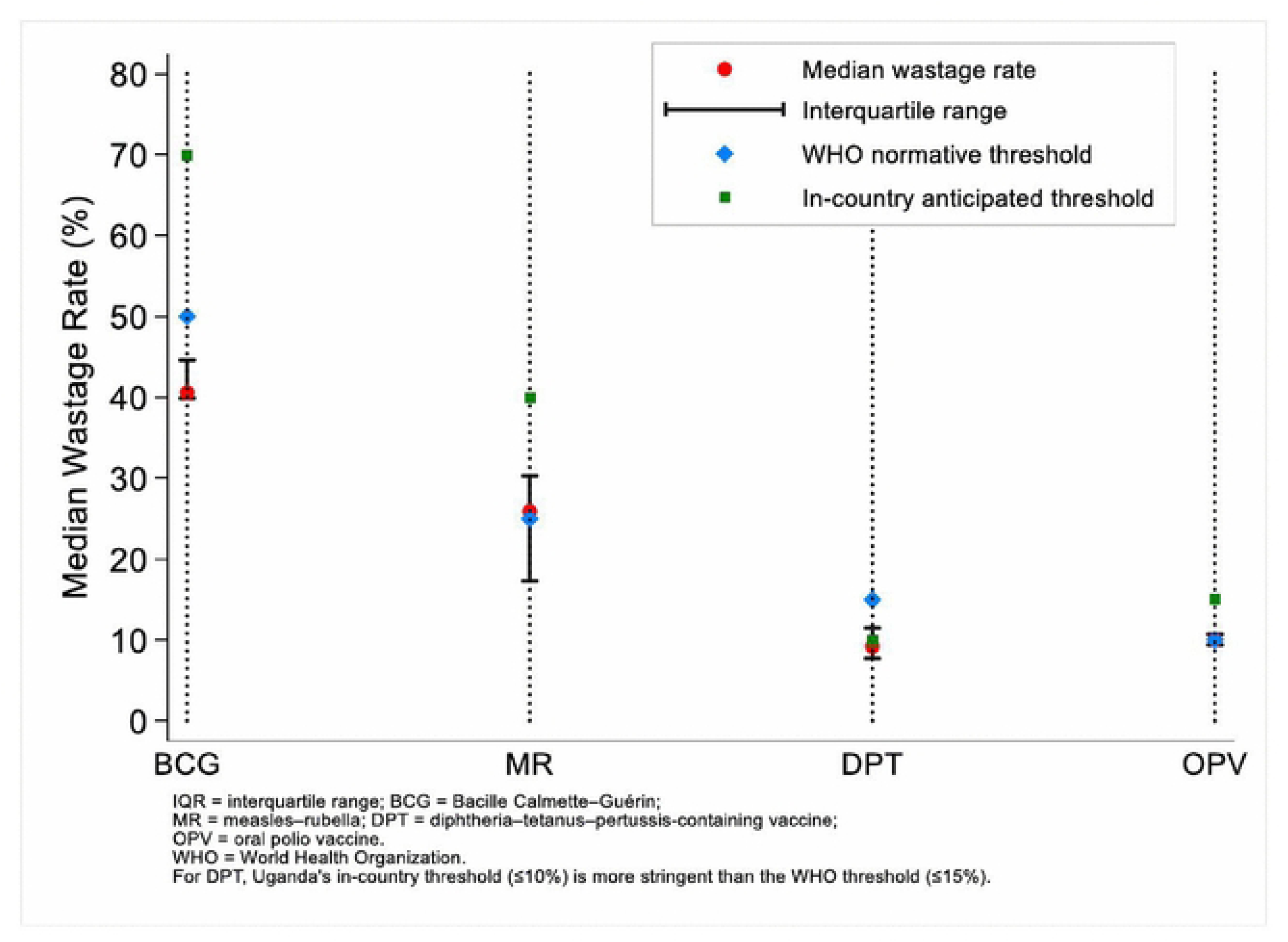
National median vaccine wastage rates compared with WHO and Uganda recommended thresholds, 2020–2025.

### Temporal trends in national vaccine wastage for the four tracer antigens, Uganda, 2020–2025

BCG wastage fluctuated markedly, with a non-significant estimated annual increase of 4.1% (95% CI −13%-**+**21%; p=0.554), and no observed monotonic trend (τ_b_=0.07; p=1.000); (Figure 2A). Similarly, MR wastage showed substantial year-to-year variation, with an estimated annual change of +0.9% (95% CI −36.7 to 38.6; p=0.95), and a non-significant monotonic trend (τ_b_=−0.07; p=1.000); (Figure 2B). A pronounced increase in BCG and MR wastage observed during 2023–2024 coincide with major programmatic events, including large-scale catch-up vaccination activities and operational adjustments following the introduction of the second dose of measles–rubella vaccine (MR2).

**Figure 2:**
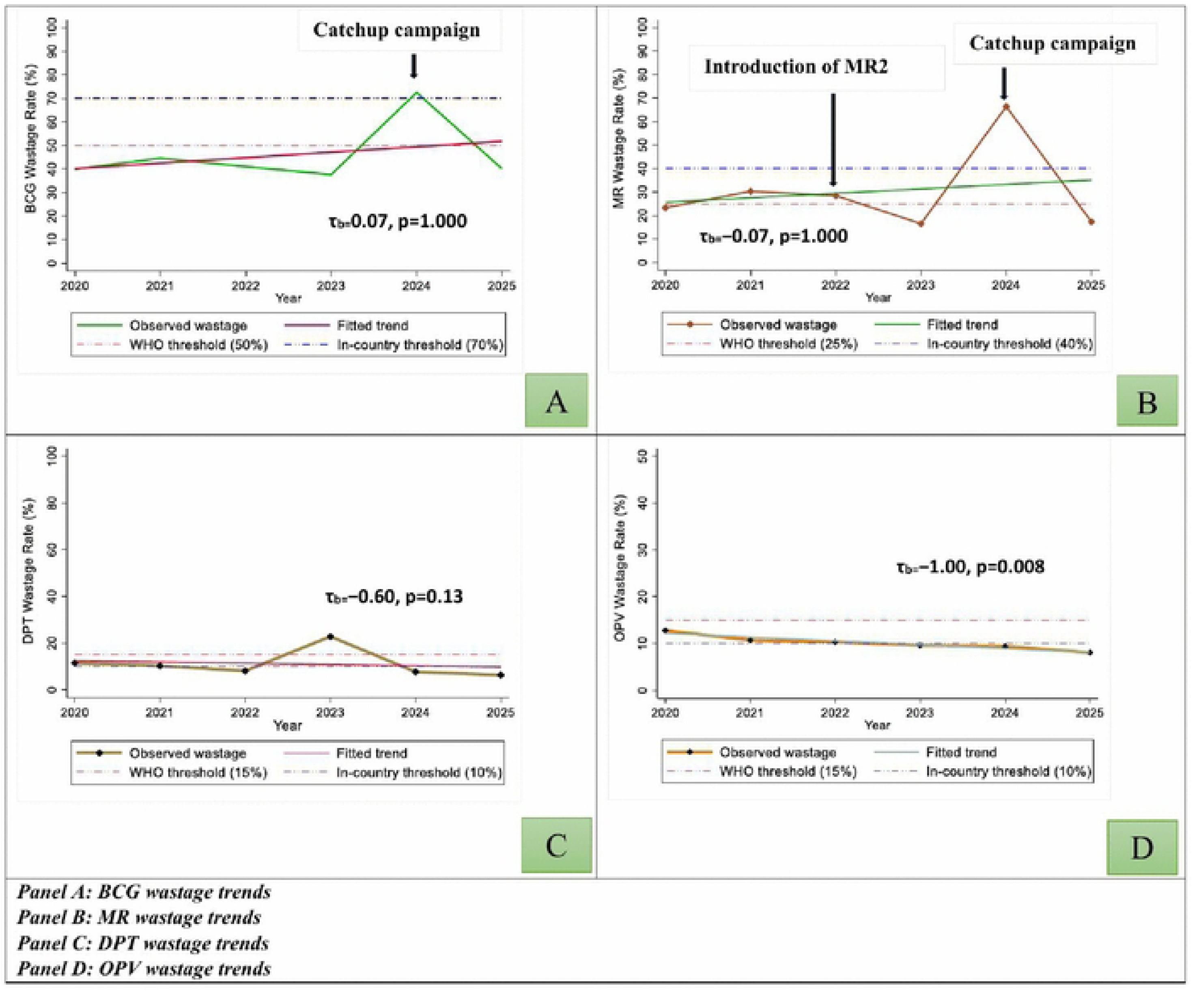
Temporal trends of National median vaccine wastage rates by antigen type, Uganda 2020-2025.

DPT wastage alternated across years, with the largest rise in 2023, before declining in 2024 and 2025; average annual reduction of −7.7% (p=0.52), with a moderate negative monotonic trend (τ_b_=−0.60; p=0.13) that didn’t reach statistical significance; (Figure 2C). In contrast, OPV wastage declined steadily with an estimated significant annual reduction of −7.6% (p=0.002) fully supported by a significant downward trend (τ_b_=−1.00; p=0.008); (Figure 2D).

### District-level threshold performance

District-level performance improved most clearly for OPV and DPT. For OPV, the proportion of districts meeting the WHO threshold of ≤10% increased from 31.5% (46/146) in 2020 to 67.1% (98/146) in 2025. Districts exceeding both WHO and Uganda thresholds declined from 36.3% (53/146) to 5.5% (8/146), showing a significant downward trend (τb=−0.87; p=0.024).

For DPT, the proportion of districts meeting Uganda’s stricter planning threshold of ≤10% increased from 50.0% (73/146) in 2020 to 88.4% (129/146) in 2025 (τb=1.00; p=0.009). Districts exceeding both WHO and Uganda thresholds declined from 22.6% (33/146) to 1.4% (2/146), also showing a significant downward trend (τb=−0.97; p=0.013).

BCG and MR showed more stable but less consistent district-level patterns. For BCG, the proportion of districts meeting the WHO threshold of ≤50% remained high, increasing slightly from 82.9% (121/146) in 2020 to 85.6% (125/146) in 2025, with no significant trend (τb=0.07; p=1.000). Districts exceeding both WHO and Uganda thresholds remained uncommon, below 3% in all years. For MR, districts meeting the WHO threshold of ≤25% increased from 63.7% (93/146) in 2020 to 82.9% (121/146) in 2025, while districts exceeding both thresholds declined from 6.8% (10/146) to 2.1% (3/146). However, the trend was not statistically significant (τb=0.33; p=0.450) (Figure 3).

**Figure 3:**
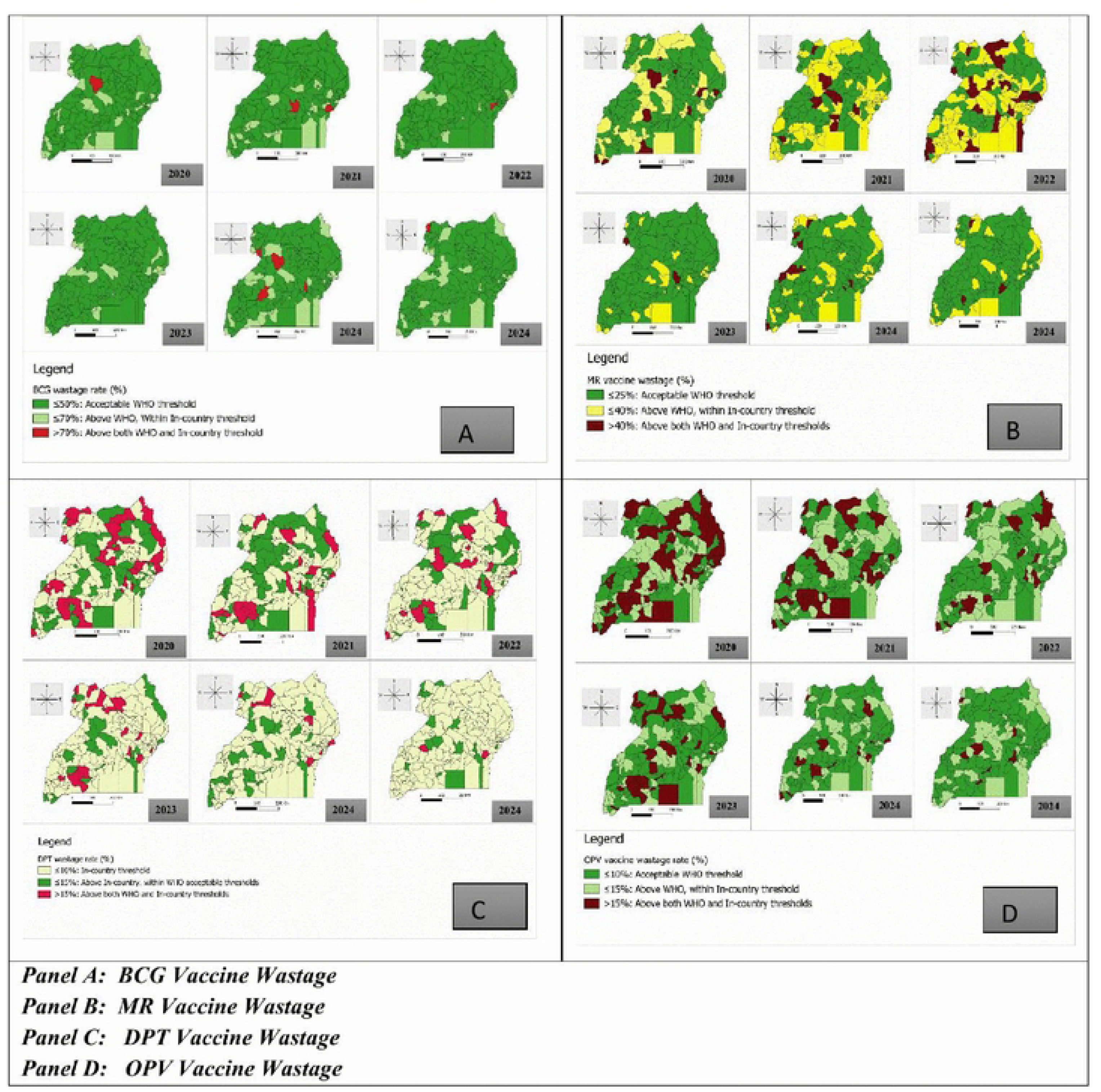
Spatial distribution of vaccine wastage compared with global and national recommended thresholds across districts in Uganda, 2020–2025.

### Vaccine wastage trends by antigen and health facility category, Uganda, 2020–2025

OPV wastage declined over time across both facility categories (Figure 4A). At lower-level facilities, wastage fell from 22.0% in 2020 to 12.9% in 2024 before rising slightly to 15.5% in 2025 (APC −9.0%, 95% CI −15.5 to −2.5; p=0.019; τ_b_=−0.86; p=0.034). Despite this decline, wastage remained above the WHO threshold (≤10%) for most of the study period and intermittently exceeded the national threshold (≤15%). Higher-level facilities showed a steeper and more sustained decline, from 14.6% to 6.0% (APC −20.9%, 95% CI −33.3 to −8.6; p=0.009; τ_b_=−0.87; p=0.024), remaining below both thresholds throughout the study period.

**Figure 4:**
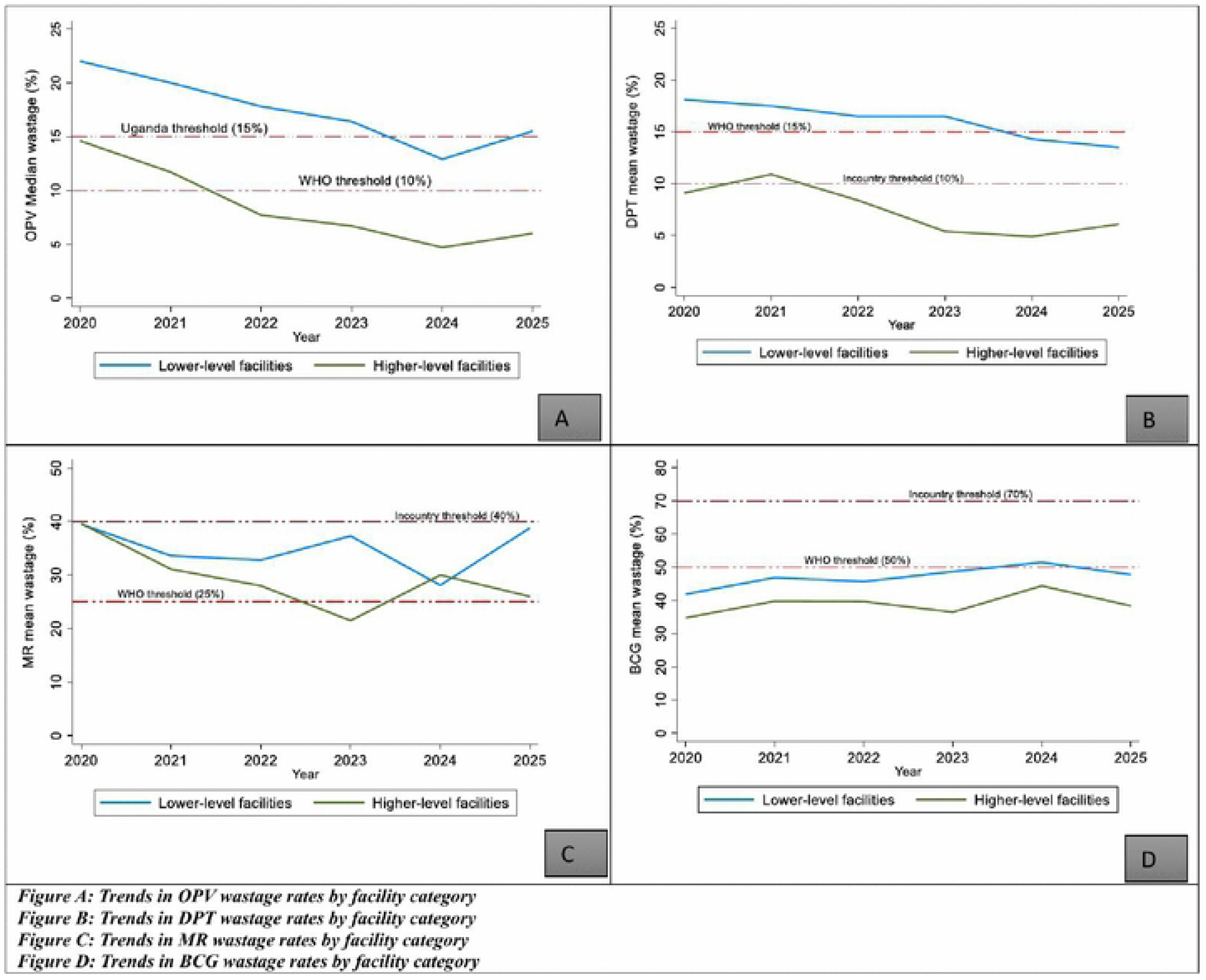
Trends of vaccine wastage rates by antigen across health facility categories in Uganda, 2020–2025.

DPT wastage was consistently higher at lower-level than higher-level facilities throughout the study period (Figure 4B). Lower-level facilities exceeded both the WHO (≤15%) and national (≤10%) thresholds for most years, though wastage declined from 18.1% in 2020 to 13.5% in 2025 (APC −5.9%, 95% CI −8.3 to −3.5; p=0.002; τ_b_=−0.97; p=0.013). Higher-level facilities maintained wastage below both WHO and national threshold in most years, with a steeper but non-significant negative trend (APC −13.8%, 95% CI −27.7 to 0.01; p=0.050; τ_b_=−0.60; p=0.133). MR wastage remained persistently high across both facility categories, frequently exceeding the WHO threshold (≤25%) while generally remaining within the national threshold (≤40%) (Figure 4C). Lower-level facilities recorded higher and more unstable wastage, fluctuating between 28.1% and 39.5%, with no significant trend (APC −1.4%, 95% CI −10.8 to 7.9; p=0.695; τ_b_=−0.20; p=0.707). Higher-level facilities showed comparatively greater improvement, declining from 39.6% in 2020 to 26.0% in 2025, though the trend did not reach statistical significance (APC −7.1%, 95% CI −18.5 to 4.3; p=0.160; τ_b_=−0.60; p=0.133).

BCG wastage remained elevated across both facility categories, frequently approaching or exceeding the WHO threshold (≤50%) while remaining below the national threshold (≤70%) throughout (Figure 4D). Lower-level facilities recorded higher wastage with a non significant upward trend, rising from 41.9% in 2020 to a peak of 51.5% in 2024 before declining to 47.8% in 2025 (APC +2.9%, 95% CI −0.4 to 6.1; p=0.070; τ_b_=0.60; p=0.133). Higher-level facilities maintained comparatively lower wastage rate, ranging from 34.8% to 44.4%, with no significant trend (APC +2.1%, 95% CI −3.4 to 7.6; p=0.347; τ_b_=0.20; p=0.707).

### Reported reasons for vaccine wastage

Findings from EVM Version 2.0 assessments and supplementary UNEPI reports conducted across 70 sites nationwide (2020 and 2025) identified vaccine wastage at multiple levels of the supply chain (Table 1).

**Table 1:**
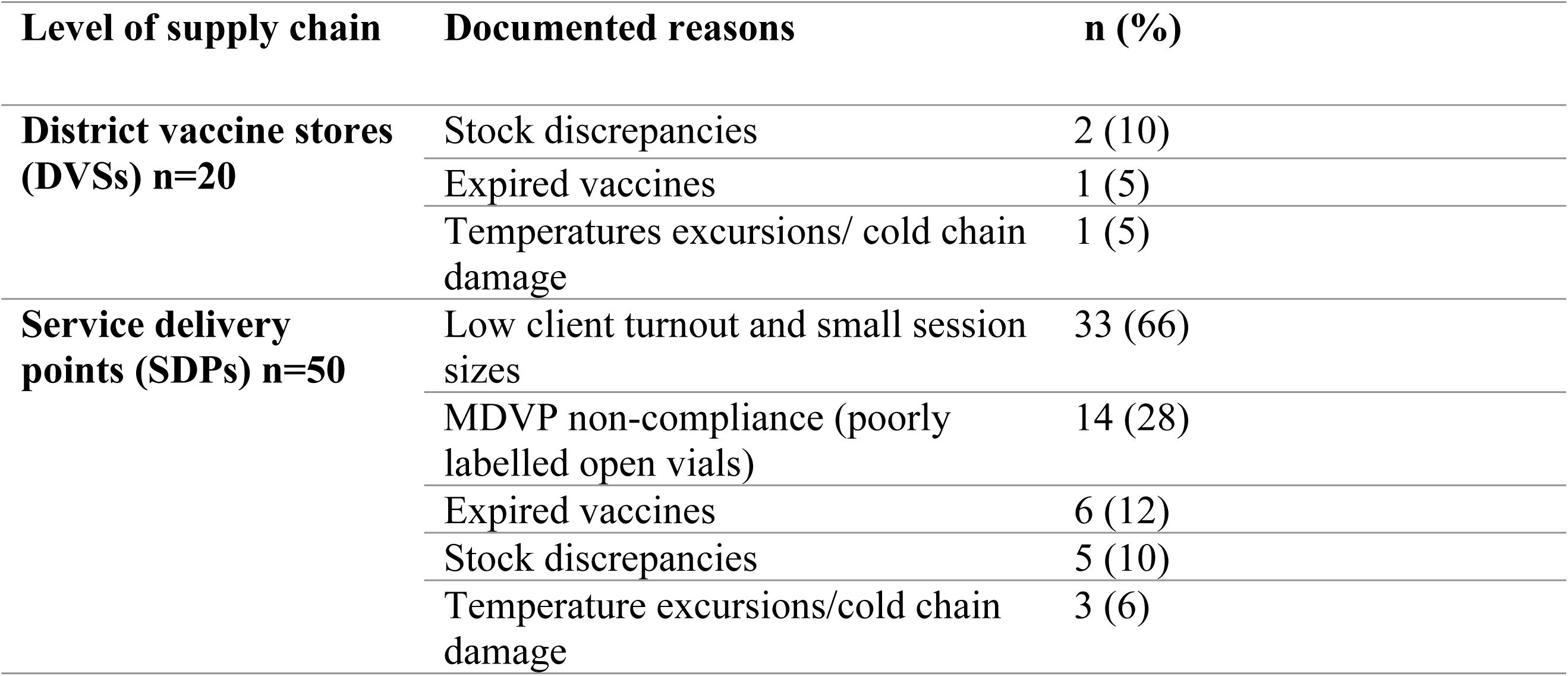
Documented reasons for vaccine wastage by supply chain level, 2020-2025 (n=70)

| Level of supply chain | Documented reasons | n (%) |
| --- | --- | --- |
| **District vaccine stores (DVSs) n=20** | Stock discrepancies | 2 (10) |
|  | Expired vaccines | 1 (5) |
|  | Temperatures excursions/ cold chain damage | 1 (5) |
| **Service delivery points (SDPs) n=50** | Low client turnout and small session sizes | 33 (66) |
|  | MDVP non-compliance (poorly labelled open vials) | 14 (28) |
|  | Expired vaccines | 6 (12) |
|  | Stock discrepancies | 5 (10) |
|  | Temperature excursions/cold chain damage | 3 (6) |

### Service Delivery Points (SDPs)

Effective Vaccine Management assessments and supplementary UNEPI reports documented vaccine wastage reasons across 70 sites, including 50 service delivery points and 20 district vaccine stores. Among the 50 service delivery points assessed, the most commonly reported reason for vaccine wastage was low client turnout or small session size, reported by 66% (33/50) of facilities. Non-compliance with multi-dose vial policy requirements, mainly poor labelling of opened vials, was reported by 28% (14/50). Vaccine expiry was reported by 12% (6/50), stock discrepancies by 10% (5/50), and temperature excursions or cold-chain-related damage by 6% (3/50).

Among the 20 district vaccine stores assessed, stock discrepancies were reported by 10% (2/20), while expiry-related losses and temperature excursions or cold-chain-related damage were each reported by 5% (1/20) of stores (Table 1).

## Discussion

This national analysis found three main patterns in vaccine wastage in Uganda during 2020–2025. First, wastage declined most clearly for OPV, and district-level threshold performance improved for both OPV and DPT. Second, BCG and MR, both lyophilised vaccines, showed greater year-to-year variability and remained more difficult to optimise. Third, wastage was consistently higher in primary care facilities (lower-level), particularly Health Centre II and III facilities, than in hospitals and referral facilities (higher-level). Together, these findings suggest that Uganda has made progress in vaccine utilisation for MDVP-eligible liquid vaccines, while wastage for lyophilised vaccines remains a persistent operational challenge linked to session size, client turnout, and vial-opening decisions.

The decline in OPV wastage and the improved district-level performance for DPT are consistent with better utilisation of liquid multi-dose vaccines. Unlike lyophilised vaccines, which must be discarded within a short period after reconstitution, preservative-containing liquid vaccines can be retained for later use if multi-dose vial policy requirements are met, including correct storage, labelling, cold-chain maintenance, and vaccine vial monitor assessment (10,18,19). However, because this was a descriptive analysis, these improvements should not be attributed solely to MDVP implementation. Other factors, including changes in service volume, stock management, supervision, reporting quality, and vaccine distribution practices, may also have contributed. The EVM findings that 28% of service delivery points had incomplete MDVP-related labelling suggest that further gains may still be possible through targeted supervision, job aids, and routine checks of opened-vial documentation (11,20).

In contrast, BCG and MR showed less consistent improvement. This pattern is programmatically plausible because both vaccines are lyophilised and must be discarded soon after reconstitution (3,8). In low-volume settings, health workers face a practical trade-off: opening a multi-dose vial for a small number of children increases wastage, while delaying vaccination to wait for more clients may risk missed opportunities and reduced timeliness. Similar challenges have been reported in settings where small session sizes and large vial presentations contributed to open-vial wastage, especially for BCG and measles-containing vaccines (20–23). The finding that low client turnout or small session size was the most commonly reported reason for wastage supports this interpretation, although these data should be viewed as contextual evidence rather than causal estimates.

The facility-level gradient is important for programme action. Higher wastage at primary care facilities should not be interpreted simply as poor performance. These facilities often operate closer to communities, serve smaller catchment populations, conduct smaller immunisation sessions, and may provide outreach services where attendance is less predictable. Facility volume, vial size, outreach delivery, and session size have been shown to influence vaccine wastage, particularly for multi-dose and lyophilised vaccines (14,15,19,24). A uniform national target may therefore mask important operational differences across facility levels. For lower-volume facilities, the most realistic goal is not to eliminate wastage, but to reduce avoidable wastage while protecting access and timely vaccination(3).

For BCG and MR, observed wastage often remained within Uganda’s planning thresholds, suggesting that current forecasting assumptions may broadly reflect service-delivery realities. For OPV and DPT, however, declining wastage and improved district-level threshold performance suggest that planning assumptions may need periodic review to avoid overestimating vaccine requirements. Such revisions should be cautious and should consider recent consumption patterns, service volume, district variability, and stockout risk (25,26).

The synchronised increase in wastage for multiple antigens during 2023–2024 may reflect system-wide operational disruptions occurring during this period. Both events typically disrupt established distribution and utilisation patterns: catch-up campaigns require pre-positioning of vaccines across multiple sites where attendance may be lower than anticipated, while new vaccine introductions are often associated with transitional wastage as quantification, training, and demand-generation systems adjust (5,20,22). For lyophilised vaccines, strict open-vial discard requirements amplify these dynamics when session sizes fall short (8,10,24).

The subsequent decline in 2025 suggests system recovery rather than sustained structural inefficiency, but the episode underscores the program’s vulnerability to operational shocks and the need for real-time monitoring systems capable of detecting abrupt deviations and enabling timely corrective action(27,28).

Finally, EVM data indicate that wastage from temperature excursions, damage, or expiry was reported in a minority of facilities, suggesting that elevated wastage, particularly for lyophilised vaccines, is more likely to be driven by open-vial losses than by infrastructure-related failures(3). Programme efforts should therefore prioritise demand aggregation and vial-size optimisation.

Persistent data quality challenges, including stock discrepancies and recording inconsistencies, nonetheless limit confidence in routine wastage estimates. Reintroducing structured reporting tools with DHIS2-integrated validation and automated outlier detection would substantially improve data reliability for programme decision-making (29).

These findings have important implications for immunisation planning and supply chain management in Uganda. The sustained decline in OPV wastage and improved DPT performance are consistent with improved utilisation of MDVP-eligible liquid vaccines, although this descriptive analysis cannot attribute these changes to specific interventions such as supervision or MDVP implementation alone. In contrast, persistently high wastage for BCG and MR at lower-level facilities may imply a structural mismatch between vial size and session demand, particularly in low-volume settings. Piloting smaller vial presentations for BCG and MR at peripheral facilities may reduce open-vial losses. Further operational research is needed to evaluate the cost-effectiveness of smaller vial sizes and to examine session-level drivers of wastage, including health worker vial-opening practices, which are not captured in routine DHIS2 data.

### Limitations

We relied on routinely reported DHIS2 data, which may contain errors or inconsistencies. Mitigation included range checks, internal consistency assessments, and triangulation with EVM findings. Annual aggregated data limited analysis of seasonal or session-level determinants, and sub-facility session-level wastage could not be assessed, where vial-opening decisions directly influence losses. Unmeasured factors, such as supervision intensity, health worker turnover, campaign activity, and community mobilisation, could confound results. Nevertheless, consistent facility-level trends and complementary EVM data support the robustness of the observed national and subnational patterns.

## Conclusion

Uganda has made meaningful progress in vaccine utilisation efficiency, particularly for MDVP-eligible liquid vaccines, while lyophilised vaccines remain a persistent challenge at lower-level facilities. Strengthening MDVP adherence through targeted supervision and job aids, and restoring standardised wastage reporting with DHIS2-integrated validation, could consolidate these gains and improve forecasting accuracy. For lyophilised vaccines, structured session microplanning at low-volume sites and cost-effectiveness analyses of smaller vial presentations represent the most actionable approaches to reducing open-vial losses.

Building real-time monitoring capacity would further protect efficiency gains against episodic operational shocks and strengthen supply chain sustainability within Uganda’s immunisation programme.

## Declarations

## Acknowledgements

We acknowledge the Ministry of Health through the Uganda Expanded Program on Immunisation (UNEPI), and the Uganda Public Health Fellowship Program for their technical guidance and data access support.

## Authors’ contributions

SN conceptualized the study, led data collection and analysis, and drafted the manuscript. CA, RM, PS YN, CL and EOO supported data analysis and technical review of the draft manuscript. CA, RM, LB and BK supported revision of the first manuscript draft. NM, YN, PS and FN contributed to tool development, and data quality assurance. CA, ARA and RM, provided technical input during the study design and data interpretation. All authors read and approved the final manuscript.

## Data availability

The datasets generated and analysed during this study are the property of the Uganda Public Health Fellowship Program and are not publicly available to protect participant confidentiality. Data can be made available from the corresponding author upon reasonable request, subject to approval by the Uganda Public Health Fellowship Program.

## Consent for publication

Not applicable.

## Ethics statements

Patient consent for publication: **Not applicable.**

## Patient and Public Involvement Statement

Not Applicable

## References

1. Sim SY, Watts E, Constenla D, Brenzel L, Patenaude BN. Return On Investment From Immunization Against 10 Pathogens In 94 Low- And Middle-Income Countries, 2011–30. Health Aff (Millwood). 2020 Aug;39(8):1343–53. doi:10.1377/hlthaff.2020.00103

2. World Health Organisation. Immunization agenda 2030: A global strategy to leave no one behind. Immun Agenda 2030. 2024 Apr 8;42:S5–14. doi:10.1016/j.vaccine.2022.11.042

3. World Health Organisation. Monitoring vaccine wastage at country level-Guidelines for program managers [Internet]. [cited 2025 Nov 13]. Available from: https://www.who.int/publications/i/item/WHO-VB-03-18-Rev-1

4. World Health Organisation. Vaccine Wastage Rates Calculator [Internet]. 2022 [cited 2025 Nov 11]. Available from: https://www.who.int/publications/m/item/vaccine-wastage-rates-calculator

5. Mvundura M, Lorenson K, Chweya A, Kigadye R, Bartholomew K, Makame M, et al. Estimating the costs of the vaccine supply chain and service delivery for selected districts in Kenya and Tanzania. Vaccine. 2015 May 28;33(23):2697–703. doi:10.1016/j.vaccine.2015.03.084

6. Lee BY, Haidari LA, Prosser W, Connor DL, Bechtel R, Dipuve A, et al. Re-designing the Mozambique vaccine supply chain to improve access to vaccines. Vaccine. 2016 Sep 22;34(41):4998–5004. doi:10.1016/j.vaccine.2016.08.036

7. World Health Organisation. Immunization Financing: A Resource Guide for Advocates, Policymakers, and Program Managers. 2016.

8. World Health Organisation. WHO policy statement: Multi-dose vial policy (MDVP) – Handling of multi-dose vaccine vials after opening [Internet]. Geneva: WHO; 2014 [Internet]. 2014 [cited 2025 Nov 11]. Available from: https://www.who.int/publications/i/item/WHO-IVB-14.07

9. World Health Organisation. UNICEF. Effective Vaccine Management (EVM) Initiative. Geneva: WHO; 2019 [Internet]. 2019 [cited 2026 Feb 17]. Available from: https://www.who.int/teams/immunization-vaccines-and-biologicals/essential-programme-on-immunization/supply-chain/effective-vaccine-management

10. Basu S, Rustagi R. Multi-dose vials versus single-dose vials for vaccination: perspectives from lower-middle income countries. Hum Vaccines Immunother. 18(6):2059310. doi:10.1080/21645515.2022.2059310 PubMed PMID: 35416750; PubMed Central PMCID: PMC9746400.

11. Wallace AS, Willis F, Nwaze E, Dieng B, Sipilanyambe N, Daniels D, et al. Vaccine wastage in Nigeria: An assessment of wastage rates and related vaccinator knowledge, attitudes and practices. Vaccine. 2017 Dec 4;35(48 Pt B):6751–8. doi:10.1016/j.vaccine.2017.09.082 PubMed PMID: 29066189; PubMed Central PMCID: PMC5771486.

12. Ministry of Health-Uganda National Expanded Programme on Immunization Multi Year Plan 2021-2025. Kampala: Ministry of Health; 2021. [Internet]. [cited 2026 Feb 16]. Available from: https://library.health.go.ug/sites/default/files/resources/UNEPI%20M%26E%20PLAN%2024-28%20final%20Version.pdf

13. Ministry of Health. The National Immunisation Strategy (NIS) 2024-2028 | MOH Knowledge Management Portal [Internet]. 2024 [cited 2025 Nov 11]. Available from: https://library.health.go.ug/communicable-disease/vaccination-and-immunisation-unepi/national-immunisation-strategy-nis-2024

14. Ninsiima M, Muhoozi M, Luzze H, Kasasa S. Vaccine wastage rates and attributed factors in rural and urban areas in Uganda: Case of Mukono and Kalungu districts. PLOS Glob Public Health. 2025 Jun 10;5(6):e0003745. doi:10.1371/journal.pgph.0003745

15. Kemigisha M, Migisha R, Kyamwanga IT. Assessment of vaccine wastage and associated factors in Rukungiri District, Southwestern Uganda, 2018–2019: a mixed-methods study. 2024.

16. UNICEF. Vaccination and Immunization Statistics - UNICEF DATA [Internet]. 2024 [cited 2026 Feb 17]. Available from: https://data.unicef.org/topic/child-health/immunization/

17. Uganda Bureau of Statistics, ICF. Uganda Demographic and Health Survey 2022. Kampala: UBOS and ICF; 2023. [Internet]. [cited 2026 Feb 16]. Available from: https://www.ubos.org/wp-content/uploads/publications/UDHS-2022-Report.pdf

18. Lydon P, Zipursky S, Tevi-Benissan C, Djingarey MH, Gbedonou P, Youssouf BO, et al. Economic benefits of keeping vaccines at ambient temperature during mass vaccination: the case of meningitis A vaccine in Chad. Bull World Health Organ. 2014 Feb 1;92(2):86–92. doi:10.2471/BLT.13.123471 PubMed PMID: 24623901; PubMed Central PMCID: PMC3949534.

19. Radwan NF, Abu-Sheasha GA, Bedwani RN, Yassine OG. Vaccine wastage and cost saving after multi-dose vial policy implementation in Egypt: A success story. Vaccine. 2021 Dec 17;39(51):7457–63. doi:10.1016/j.vaccine.2021.10.067

20. Mvundura M, Ng J, Reynolds K, Theng Ng Y, Bawa J, Bambo M, et al. Vaccine wastage in Ghana, Mozambique, and Pakistan: An assessment of wastage rates for four vaccines and the context, causes, drivers, and knowledge, attitudes and practices for vaccine wastage. Vaccine. 2023 Jun 23;41(28):4158–69. doi:10.1016/j.vaccine.2023.05.033

21. Setia S, Mainzer H, Washington ML, Coil G, Snyder R, Weniger BG. Frequency and causes of vaccine wastage. Vaccine. 2002;20(7–8):1148–56.

22. Bawa S, Shuaib F, Saidu M, Ningi A, Abdullahi S, Abba B, et al. Conduct of vaccination in hard-to-reach areas to address potential polio reservoir areas, 2014–2015. BMC Public Health. 2018 Dec 13;18(Suppl 4):1312. doi:10.1186/s12889-018-6194-y PubMed PMID: 30541501; PubMed Central PMCID: PMC6291919.

23. GAVI. Gavi, the Vaccine Alliance. Supply Chain Strategy. Geneva: Gavi; 2020. [Internet]. 2020 [cited 2026 Feb 25]. Available from: https://www.gavi.org/our-work/health-systems-immunisation-strengthening/immunisation-supply

24. Parmar D, Baruwa EM, Zuber P, Kone S. Impact of wastage on single and multi-dose vaccine vials: Implications for introducing pneumococcal vaccines in developing countries. Hum Vaccin. 2010 Mar;6(3):270–8. doi:10.4161/hv.6.3.10397

25. Lydon P, Schreiber B, Gasca A, Dumolard L, Urfer D, Senouci K. Vaccine stockouts around the world: Are essential vaccines always available when needed? Vaccine. 2017 Apr 19;Building Next Generation Immunization Supply Chains35(17):2121–6. doi:10.1016/j.vaccine.2016.12.071

26. Oo AN, Thekkur P, Thar AMC, Htet KKK, Lin HH. Small Session Size and Big Vial Size: Operational Research Assessing Open Vial Vaccine Wastage at the Service Delivery Points in the Mandalay Region of Myanmar During 2018. Trop Med Infect Dis. 2020 Apr 15;5(2):60. doi:10.3390/tropicalmed5020060 PubMed PMID: 32326568; PubMed Central PMCID: PMC7344912.

27. World Health Organisation. Progresses and Challenges with Sustaining and Advancing Immunization Coverage During the COVID-19 Pandemic [Internet]. 2023 [cited 2025 Nov 8]. Available from: https://www.who.int/publications/i/item/progresses-and-challenges-with-sustaining-and-advancing-immunization-coverage-during-the-covid-19-pandemic

28. Shet A, Carr K, Danovaro-Holliday MC, Sodha SV, Prosperi C, Wunderlich J, et al. Impact of the SARS-CoV-2 pandemic on routine immunisation services: evidence of disruption and recovery from 170 countries and territories. Lancet Glob Health. 2022 Feb 1;10(2):e186–94. doi:10.1016/S2214-109X(21)00512-X PubMed PMID: 34951973.

29. Lim J, Norman BA, Rajgopal J. Redesign of vaccine distribution networks. Int Trans Oper Res. 2022;29(1):200–25.

